# Multicenter analysis of immunosuppressive medications on the subsequent risk of malignancy in solid organ transplant recipients

**DOI:** 10.1101/2022.12.02.22283027

**Authors:** Reid Shaw, Ali R. Haque, Jack Fitzsimons, Adam Hamidi, Timothy E. O’Connor, Gregory W. Roloff, Bradford C. Bemiss, Eric R. Kallwitz, Patrick A. Hagen, Stephanie Berg

## Abstract

Solid organ transplant is a curative treatment for end organ disease. However, the immunosuppressive therapy required to prevent graft rejection increases the likelihood of developing a subsequent malignancy. This retrospective cohort study from a multi-center academic hospital system investigated solid organ transplantation, immunosuppression, and the risk of subsequent malignancy. Of the 5,591 patients and 6,142 transplanted organs studied, there were 517 subsequent malignancies identified. Skin cancer was the most common type of malignancy to be diagnosed, whereas liver cancer was the first malignancy to present at a median time of one-year post-transplant. Subsequent malignancy was proportionally more often diagnosed in non-Hispanic White transplant recipients compared to other racial groups. Heart and lung transplant recipients had relatively higher rates of subsequent malignancy than liver and kidney transplant recipients, but this finding was not significant upon adjusting for immunosuppressive medications. Multivariate cox proportional hazard analysis and random forest variable importance calculations identified statistically significant correlations with sirolimus and azathioprine and high rates of malignancy after transplant, while tacrolimus was associated with low rates of post-transplant neoplasia.

## 1 Introduction

Solid organ transplant (SOT) is a curative treatment option for many patients with end-stage organ disease.^1^ In 2021, there were more than 13,800 deceased and 6,500 living donor transplants in the United States.^2^ Although transplant-related outcomes have significantly improved over time, rates of morbidity and mortality after successful transplantation represent areas for clinical improvement.^3^ One major adverse outcome after SOT is malignancy with standardized incidence ratios of 2-4 times that of the general public.^4,5^ This is due to a variety of patient,^6,7^ donor,^8^ transplant,^9^ and medication-related factors.^8,10^

Immunosuppressive therapy is considered to be a significant risk factor in the development of a malignancy following SOT as it may lead to the activation of oncogenic viruses^11^, dysfunction of DNA repair^12^, and other immune-mediated mechanisms.^13^ Prior studies have assessed the risk of malignancy across different organ types of transplantation, while others have investigated immunosuppressive regimens on the risk of malignancy. However, no study has assessed the risk of malignancy across all organ types with respect to immunosuppression. This study aims to identify potential risk factors associated with developing malignancy across all SOT types. Understanding the malignancy risks associated with immunosuppressive medications across all organ transplant types may lead to better informed patients and clinicians, allowing for the maintenance of graft function while limiting associated morbidity and mortality.

## 2 Methods

### 2.1 Study design and data collection

This is an IRB-approved, retrospective cohort study from three academic hospitals in the greater Chicago area Loyola University Medical Center, Gottlieb Memorial Hospital, and MacNeal Hospital. The electronic health record (EHR) software (Epic Systems; Verona, WI) was queried from January 1, 2000 to March 10, 2021. SOT and malignancies were identified using a complete list of international classification of diseases (ICD) codes from the 9^th^ and 10^th^ revision. The date of SOT and diagnosis of malignancy was defined as the first instance an ICD code appeared in a patient’s medical record. To ease in subsequent analysis, similar ICD-9 and ICD-10 diagnoses were grouped together into nominal variables. The histological evaluation of each malignancy was not captured within the ICD codes of this dataset. A complete list of medications and patient demographics, including age, sex, race, ethnicity, zip-code, and preferred language were also queried from the EHR. Patients under the age of 18 were not included in this study.

### 2.2 Statistics

A two-sided t-test was calculated to assess differences in numerical variables. Chi-squared was used to assess differences in proportions. Pearson’s was used to assess for correlation between dichotomous variables. Loess smoothing was used to assess trends overtime.^14^ For time-to-event analysis, patients were censored when diagnosed with a malignancy, or the last time a medication was taken, representing the last time of contact with the study center or death. For each patient, every transplant and new malignancy diagnosis represented a new observation within the data frame. Gray’s test was used for the competing risk analysis.^15^ Immunosuppressive medications were required to be taken for at least one day. Multivariable regression was performed using Cox proportional hazard with time to malignancy as the dependent variable. Independent variables included age at transplant, race, sex, immunosuppressive medications, and transplanted organ. Statistical significance was defined as p-value < 0.05 and was adjusted with a Bonferroni correction.^16^

### 2.3 Machine learning

Variable importance calculations were performed as previously described.^17^ Categorical variables with a frequency of less than 10% were grouped into an “other” category. Nominal variables were one-hot encoded and numerical variables were normalized to have a standard deviation of one with a mean of zero. The complete dataset was then bootstrap resampled 10 times and stratified by subsequent malignancy. A random forest model was then fit across a variety of hyperparameters within a Latin hypercube of size 25.^18^ Gini impurity values were calculated to provide robust assessment of variable importance.^19^

### 2.4 Analysis code

All analysis was performed with R programming language v.4.0.3.^20^ using the ‘tidyverse,’^21^ ‘tidymodels,’^22^ ‘lubridate,’^23^ ‘funneljoin,’^24^ ‘ggridges,’^25^ ‘surival,’^26^ ‘janitor,’^27^ ‘wesanderson,’^28^ ‘ochRe,’^29^ ‘scales,’^30^ ‘vip,’^31^ ‘resample,’^32^ ‘corrplot,’^33^ and ‘tidycmprsk’^34^ packages.

## 3 Results

### 3.1 Transplant recipient descriptive statistics

Overall, there were 5,591 unique patients who received a SOT during the queried time period, comprising six different organ types and 6,142 transplanted organs (**Figure 1A**). Kidney transplants (n = 2,986) were the most common, followed by liver (n = 1,298), lung (n = 1,024), heart (n = 723), pancreas (n = 106), and intestine (n = 5) (**Figure 1A**). The median age of a transplant recipient was 54 years with a range of 18-91 years (**Supplementary Figure 1A**). 5,093 people received one SOT, 448 received two organs, 47 received three organs, and three received four organs (**Supplementary Figure 1B**). 64% of individuals to receive a transplant self-identified as White, while the next most common racial demographic was Black at 17% (**Supplementary Figure 1C**). White and Asian transplant recipients were the oldest demographic to receive an organ at a median age of 56 years (**Figure 1B**). These two groups were statistically older than other minority groups, including Black (52 years), Hispanic (50 years), and multi-racial (40 years) (**Figure 1B**). The median age of transplantation was highest in lung (58 years), heart (58 years), and liver (59 years) transplantation (**Figure 1C**). Overall, 39% of transplant recipients were women. With the exception of intestine, SOT was more common in men than in women (**Figure 1D**). Since the beginning of this study, the number of SOT across time was increasing (**Figure 1E**). Loess smoothing showed a trend towards women becoming less likely to receive SOT when compared to men (**Figure 1F**).

**Figure 1.**
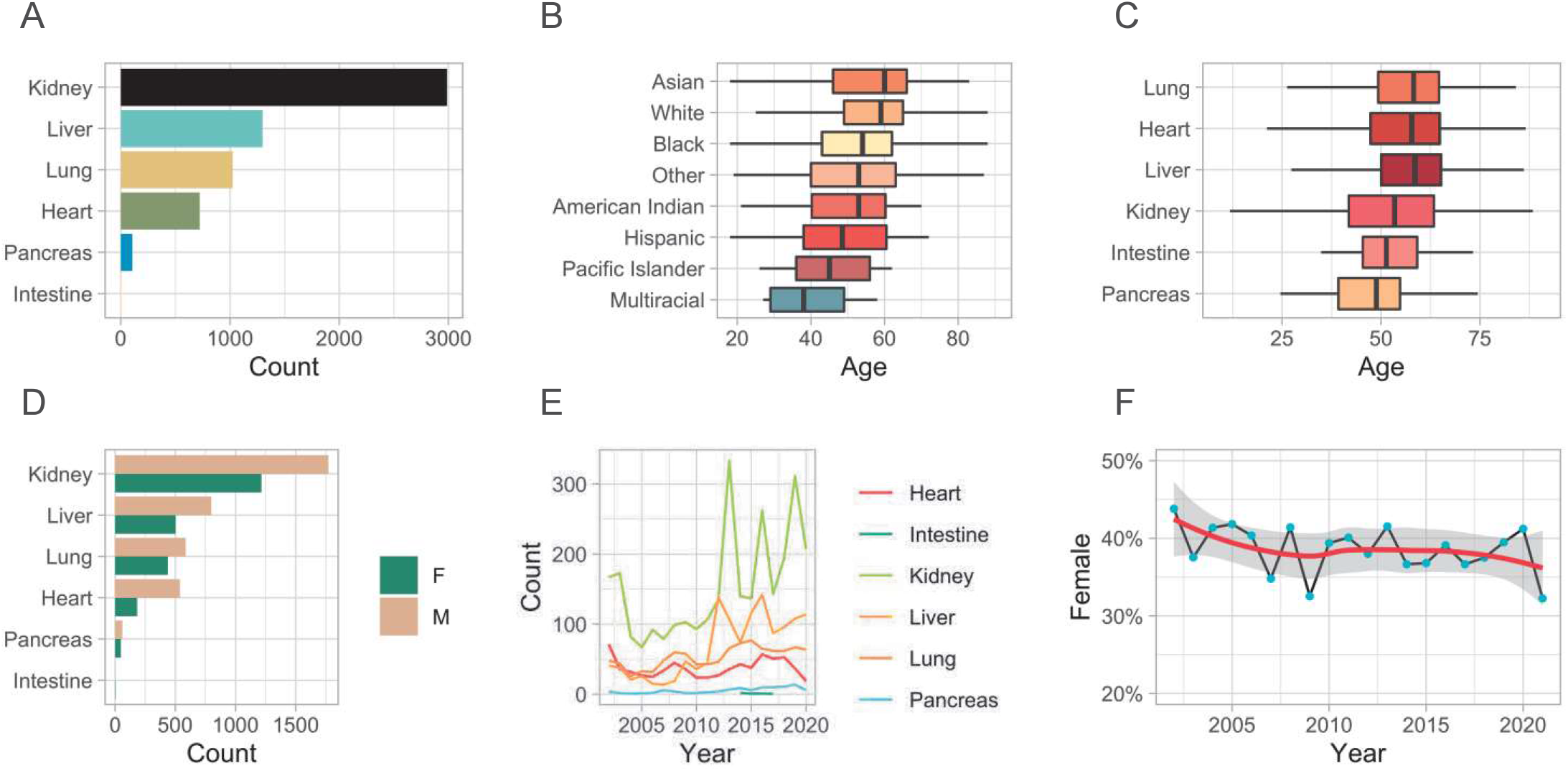
A) Bar plot of the overall number of solid organ transplants stratified by organ type. B) Boxplots of the age of solid organ transplantat stratified by race C) Boxplots of the age of solid organ transplant grouped by organ type. D) Grouped bar plot of solid organ transplant types stratified by sex. Male is denoted by ‘M’ and Female is denoted by ‘F’. E) Solid organ transplant recipients across time stratified by organ type. F) Dot plot showing the percent of women who received a solid organ transplant across the study time period. A loess smoothed line is overlayed.

### 3.2 Subsequent malignancy descriptive statistics

Of those that received a SOT, there were 517 (8.25%) malignancies identified after transplantation. Skin cancer was the predominant malignancy across SOT recipients (n = 273), followed by lymphoma (n = 40), and kidney (n = 30) (**Figure 2A**). In liver and kidney transplant recipients, subsequent liver and kidney malignancies represent the largest proportional increase from baseline rates, respectively (**Figure 2A**). Liver cancer was the earliest malignancy to present following SOT at an average of 351 days (**Figure 2B**). The median presentation of skin cancer was 1,073 days, breast cancer at 1,109 days, and lymphoma at 1,123 days (**Figure 2B**). Leukemia (n = 12) was the malignancy with the longest post-transplant latency time to presentation at just under five years (1,735 days) (**Figure 2B**). 14% of White transplant recipients were diagnosed with a malignancy after SOT; Hispanic and Black individuals were diagnosed at 9% and 7%, respectively (**Figure 2C**) Of the 249 Asian patients who received SOT, only four (1.6%) developed a subsequent malignancy (**Figure 2C**). Lung transplant recipients who developed a subsequent malignancy were significantly younger than those who did not (p = 0.048) (**Figure 2D**). No other SOT recipient group had a significant difference in subsequent malignancies based on the age of transplantation (**Figure 2D**).

**Figure 2.**
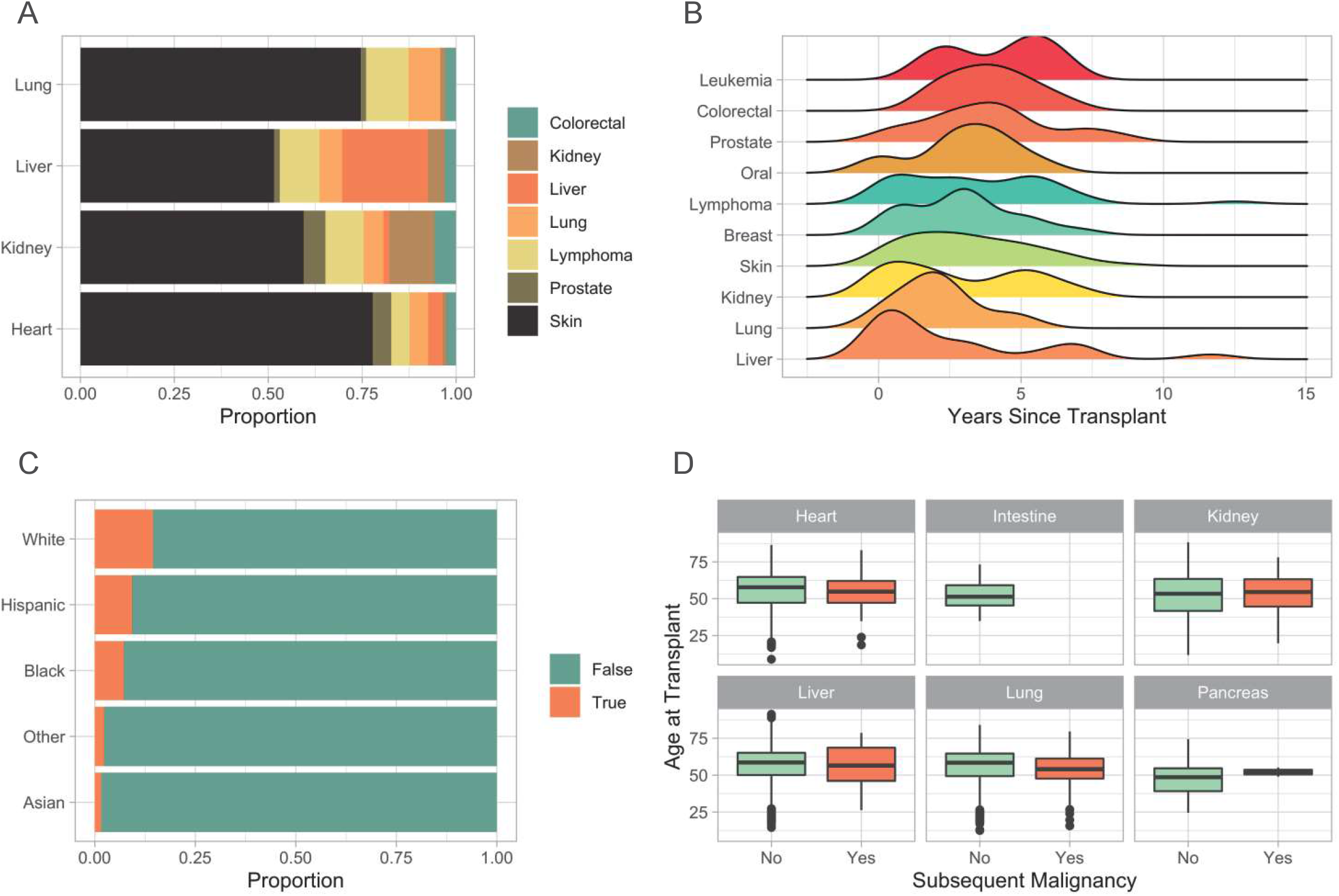
A) Stacked bar plot of subsequent malignancies by solid organ transplant type. B) Ridge plot of subsequent malignancies from time of transplant. C) Stacked bar plot of the proportion of subsequent malignancies stratified by race. True indicates the development of a subsequent malignancy. D) Boxplots of subsequent malignancies by age at transplant stratified by organ type.

### 3.3 Immunosuppression and development of malignancy

The most common immunosuppressive medications used were mycophenolate, and tacrolimus (**Supplementary Figure 2A**). The use of these medications increased throughout the study duration (**Supplementary Figure 2B**). Cyclosporine and sirolimus represent the largest proportion of immunosuppressive medications utilized in heart transplant recipients, at 12% and 10%, respectively (**Figure 3A**). Azathioprine was most commonly used in lung transplant recipients at 33% and 91% of liver transplant recipients received tacrolimus (**Figure 3A**). Tacrolimus and mycophenolate (phi coefficient 0.36) were the drugs most commonly used in the same patients, whereas tacrolimus and cyclosporine (phi coefficient -0.36) were most rarely used together (**Supplementary Figure 3**). Upon fitting a random forest machine learning (ML) model to assess variable importance on the development of malignancy, age at transplantation was the most predictive variable followed by the total number of immunosuppressive medications a patient has received (**Figure 3B**). The immunosuppressive medication with the strongest association with post-transplant malignancy was sirolimus, followed by azathioprine and tacrolimus (**Figure 3B**).

**Figure 3.**
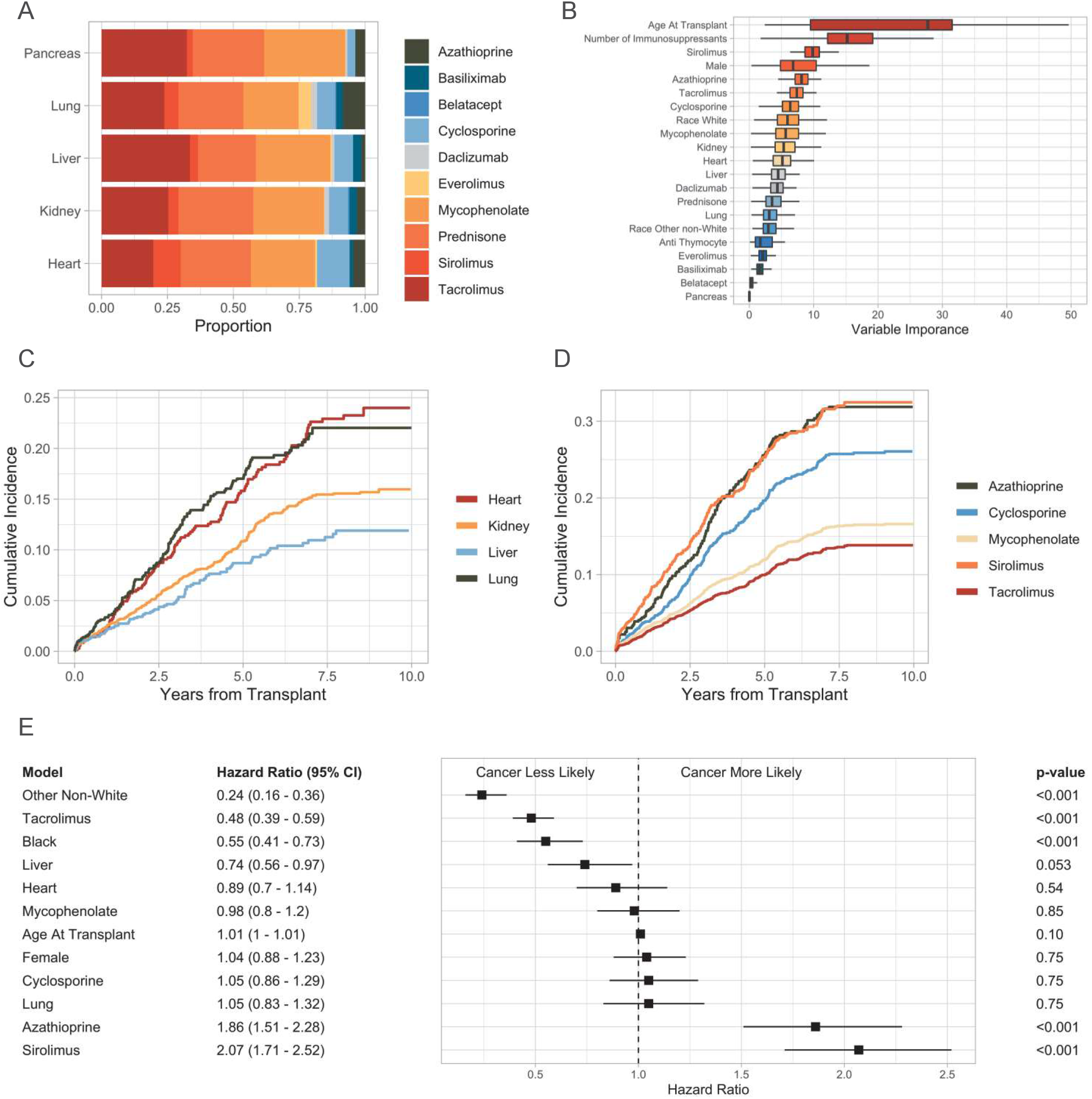
A) Stacked bar plot of immunosuppressive medications stratified by organ type. B) Boxplots of random forest Gini impurity values indicating variable importance in predicting malignancy following solid organ transplant. C) Subsequent malignancy cumulative incidence plot stratified by organ type. D) Subsequent malignancy cumulative incidence plot stratified by immunosuppressive medication. E) Forest plot of the multivariate Cox proportional hazard ratios. Black and Other Non-White hazard ratios are in comparison to White. Heart, liver, and lung are in comparison to kidney transplant recipients. P-values are adjusted with a Bonferroni correction factor.

Heart and lung transplant recipients have a high cumulative incidence of subsequent malignancy in our study (**Figure 3C**). Within 30 months post-transplant, 8.5% of heart and 8.7% of lung transplant recipients had a subsequent diagnosis of cancer (**Supplementary Figure 4**). At five years, this number increased to 16% and 17% for heart and lung transplant recipients, respectively (**Supplementary Figure 4**). Cumulative incidence of subsequent malignancy was greatest in patients who received sirolimus, azathioprine, and cyclosporine, whereas mycophenolate, and tacrolimus had a relatively lower cumulative incidence (**Figure 3D**). Multivariate Cox proportional hazard analysis demonstrated an increased risk of malignancy with azathioprine (HR 1.86, 95% CI 1.51 – 2.28, p < 0.001), and sirolimus (HR 2.07, 95% CI 1.71 – 2.52, p < 0.001), whereas patients who receive tacrolimus (HR 0.48, 95% CI 0.39 0.59, p < 0.001) were less likely to be diagnosed with a subsequent malignancy (**Figure 3E**). The differences seen in the cumulative incidence analysis stratified by organ type did not persist when adjusted for multiple variables, such as race and immunosuppressive medications (**Figure 3E**). There was no significant difference in the risk of subsequent malignancy between men and women or the age in which someone received a SOT (**Figure 3E**). Black (HR 0.55, 95% CI 0.41 0.73, p < 0.001) and other non-White (HR 0.24, 95% CI 0.16 0.36, p < 0.001) SOT recipients were less likely to develop a malignancy than White individuals (**Figure 3E**). However, there was no difference in the rates of non-cutaneous malignancies between Black and White transplants recipients (**Supplementary Figure 5**).

## 4 Discussion

To our knowledge, this is the first study to investigate the role of immunosuppression in the development of malignancy following transplantation across multiple organ types of transplantation. Our data are consistent with other landmark studies. For example, skin cancer was the most common malignancy diagnosed after SOT in our cohort, followed by lymphoma and kidney cancer.^35^ In concordance with the largest study of cancer risk in SOT recipients, our data demonstrated that the majority of liver cancer diagnoses occur within the first year of transplantation and kidney cancer incidence was highest in kidney transplant recipients.^5^

A recent population-based cohort study in Finland found increased incidence of cancer rates in heart and lung transplant recipients in comparison to kidney and liver transplant recipients.^35^ However, upon adjusting for immunosuppressive medications, we found no difference in the rates of malignancy between SOT groups, indicating the increased risk may be due to the medication. In liver transplant recipients, cumulative exposure to tacrolimus increased the risk of cancer.^36^ This finding was not unsurprising and not in opposition to our data as we did not assess cumulative exposure to tacrolimus. However, our data demonstrated a lower risk of cancer in individuals who receive tacrolimus compared to other immunosuppressive medications.

In our study, azathioprine and sirolimus were associated with the highest risk of cancer development. Azathioprine, an antagonist of purine metabolism, has long been associated with the development of cancer in SOT, inflammatory bowel disease, multiple sclerosis, and rheumatoid arthritis.^37-41^ However, the increased risk associated with sirolimus was unexpected as prior studies have generally demonstrated decreased risk of cancer with sirolimus use.^42,43^ In kidney and heart transplant recipients, the transition from a calcineurin inhibitor to sirolimus was associated with a lower risk of malignancy,^44-46^ likely due in part to the role mTOR plays in cell proliferation.^47^

ML algorithms are now being applied to a variety of SOT research questions.^48^ To our knowledge, our study represents the first time that a ML model has been used to assess variable importance in determining which SOT recipients developed a malignancy. Random forest classification, a form of decision trees, is a highly flexible, interpretable, and accurate method of estimating non-linear relationships – an area where traditional statistics struggle.^49^ In contrast to traditional statistical methods used in this analysis, our ML model identified age at transplant as a highly predictive marker of subsequent malignancy diagnosis. However, not all results were dissimilar, as the ML model also identified sirolimus, tacrolimus, azathioprine, and cyclosporine as highly predictive variables.

There are a number of limitations to this study. First, there are likely discrepancies in data entry, collection, and classification that may exist as this was a retrospective cohort study based on ICD codes. The rates of malignancy are likely underestimated as we included individuals who received transplants up to the study endpoint, and patients may be lost to follow-up for a variety of reasons. In addition, the hospitals queried in this study are located in the greater Chicago area, and thus may not represent results from distinct geographical regions across the United States or in other countries. Furthermore, we did not account for the dose of immunosuppression or blood plasma level of these medications. We also did not control for any pre-transplant related criteria, including organ ischemic time, viral studies, donor information, screening tests, or education level. This may confound some of the findings that we attribute to sociodemographic factors and immunosuppressive medications. Lastly, our findings do not establish causality. Further work will be focused on parsing the complex relationship of concomitant immunosuppression and medication changes that may affect risk of malignancy. With greater numbers of SOT recipients, we may be able to associate subsequent malignancy type with individual immunosuppressive regimens, allowing for personalized cancer screening recommendations with the goal of improving overall survival.^50^

## Data Availability

All data produced in the present study are available not currently available.

## Abbreviations

SOT: solid organ transplant
EHR: electronic health record
ICD: international classification of disease
HR: hazard ratio
CI: confidence interval
ML: machine learning
mTOR: mammalian target of rapamycin

## Acknowledgements

Many thanks to Susan Zelisko for querying the data from the EHR for subsequent analysis.

## Funding

No funding sources were used for this research project.

## Disclosures

The authors of this manuscript have no conflicts of interest to disclose as described by the *American Journal of Transplantation*.

## Notes

### Competing Interest Statement

The authors have declared no competing interest.

### Funding Statement

This study did not receive any funding.

### Author Declarations

IRB exempt per Loyola University Medical Center IRB committee.

